# Political Economy Analysis of Health Taxes (Tobacco, Alcohol Drink, and Sugar Sweteened Beverage): Case Study of Three Provinces in Indonesia

**DOI:** 10.1101/2024.01.31.24302078

**Authors:** Abdillah Ahsan, Nadira Amalia, Krisna Puji Rahmayanti, Nadhila Adani, Nur Hadi Wiyono, Althof Endawansa, Maulida Gadis Utami, Adela Miranti Yuniar, Erika Valentina Anastasia, Yuyu Buono Ayuning Pertiwi

## Abstract

Efforts to implement health tax policies to control the consumption of harmful commodities and enhance public health outcomes have garnered substantial recognition globally. However, their successful adoption remains a complex endeavour. This study takes a problem-driven political economy analysis (PEA) approach to investigate the challenges and opportunities surrounding health tax implementation, with a particular focus on sub-national government in Indonesia, where the decentralization context of health tax remains understudied. Employing a qualitative methodology by collecting data from a total of twelve focus group discussions (FGDs) conducted in three provinces—Lampung, Special Region of/*Daerah Istimewa* (DI) Yogyakarta, and Bali, each chosen to represent a specific commodity: tobacco, sugar-sweetened beverages (SSBs), and alcoholic beverages—we explore the multifaceted dynamics of health tax policies. These FGDs involved 117 participants, representing governmental institutions, non-governmental organizations (NGOs), and consumers. Our findings reveal that while health tax policies have the potential to contribute significantly to public health, challenges such as a lack of consumer awareness, bureaucratic complexities, and decentralized governance hinder implementation. Furthermore, this study underscores the importance of effective policy communication. It highlights the importance of earmarking health tax revenues for public health initiatives. It also reinforces the need to see health taxes as one intervention as part of a comprehensive public health approach including complementary non-fiscal measures like advertising restrictions and standardized packaging. Addressing these challenges is critical for realizing the full potential of health tax policies.

## Introduction

The implementation of health taxes has been successfully adopted in numerous countries and is a pivotal policy instrument to control the consumption of harmful commodities and improve public health outcomes(1-5). This approach has been endorsed as one of the “best buys” by the World Health Organization (WHO) for addressing Non-Communicable Diseases (NCDs) (6–9). Despite its importance, many countries, particularly low- and middle-income economies, still struggle to adopt this policy(10). For this reason, global health leaders recently called attention to the urgent need for research that sheds light on the politics of health tax policy processes(11).

The political economy of a health system shapes policy development and implementation in important ways(12). Conflicts of interests, poor interagency and multisectoral coordination, as well as excessive bureaucracy were among the most common challenges to fully institutionalizing evidence-informed health taxes policy recommendations(13–15). Public sector decentralization may hinder the health policy adoption due to challenges including limited shared understanding of health tax objectives among local authorities, political complexities, and financing issues at the sub-national level, resulting in suboptimal implementation (14, 16). Indonesia, as one of the largest countries with escalating concerns in the healthcare sector, primarily due to rising the NCDs epidemic(17–19). The country faces challenges in health policy adoption, primarily attributed to political contestation, industry interference, and a rigid bureaucratic system, exacerbated by decentralization(20–23).

### The current status of health tax implementation in Indonesia

Indonesia has been decentralizing since 2001—nearly 56 years after its independence(24). One crucial aspect of decentralization is public finance, encompassing various fiscal policy authorities, such as revenue generation, earmarking, and spending mandates, which has recently undergone reforms through Law No. 1/2022 (25, 26). Regarding health tax policy, both tobacco and alcoholic beverage taxes have been designed to involve sub-national governments(27, 28). Specifically for the tobacco tax collected through the Ministry of Finance, i.e., excise tax, the national government also enacted legislation to establish a tobacco-tax sharing fund (*Dana Bagi Hasil Cukai Hasil Tembakau/*DBHCHT), with the purpose of allocation to sub-national leaders based on their levels of tobacco farming production(29).

Table provides a summary of the division of roles between the national and sub-national governments in Indonesia. There are many types of taxes for commodities that fall under ‘unhealthy’ products category(30): (i) Excise Tax, regulated under Law No. 39/2007, in conjunction with Law No. 7/2021; (ii) Local Tax, which currently exclusively applies to tobacco products (as part of other five categories outlined in Law No. 28/2009, which have been subsequently replaced by Law No. 1/2022); and (iii) Value-Added Tax. In Indonesian case, health tax is more in line with excise tax, as one of its primary objectives is consumption control—rather than solely revenue generation.

Currently, only two commodities are subject to health tax: tobacco and alcoholic beverages. Although sugar-sweetened beverages (SSBs) were proposed in 2019, these have not yet been implemented(31). The national government primarily focuses on tax collection, while the sub-national government plays a critical role in tax management and allocation. Additionally, regulations mandate that sub-national governments allocate health taxes for the healthcare sector, encompassing both promotive and preventive measures(20, 32).

With the current practice of health taxation, criticisms have emerged. The health tax initiative relies heavily on the national government with the Ministry of Finance (MoF) and Ministry of Health (MoH) as policy champions(23). The reliance on the national government may be exacerbated by a limited understanding of the objectives of health taxation and complicated politics implement, particularly among sub-national governments (22, 33). The current practice prioritizes revenue generation over consumption control, especially for sub-national leaders who aim to protect revenues (20). Finally, decentralization continues to face several issues, with one of the most significant being financing despite regulations mandating health tax allocation for this sector(18, 34, 35). The financing issues include budget insufficiency and sub-optimal public insurance coverage due to disparities in revenue generation across region(35).

While decentralization becomes central to policy implementation, understanding the sub-national government behaviour in response to health tax remains important, yet understudied. This study fills this gap by exploring the political economy of health tax policy implementation at sub-national level. We selected three provinces to represent three different taxable commodities: tobacco, alcoholic beverages, and SSBs. The rationale for choosing these diverse commodities as the focus of the study is to investigate the political economy in light of their distinct attributes and characteristics. Furthermore, our focus is exclusively on the sub-national level, as there is an abundance of evidence regarding the political economy of health taxation in Indonesia at the national level (20, 21, 23, 36–39). The next section will describe the methodology and analytical approach used in this study, followed by results, and finally discussion and future recommendation.

## Material and methods

### Study design and case selection

This study is qualitative. We used an extensive case study approach as the information to be gathered requires in-depth insights and is highly contextual(40). A case study is also more suitable to answer the “why” and “how” questions which make them more explanatory and descriptive(41), aligning it with the objectives of this research.

The sub-national context is documented at the provincial level, which represents the highest tier of local government. We choose the provincial level government because it plays an important role in communicating the central government health policy while simultaneously hold a discretion over districts’ budgeting process(42). Each province was selected based on their level of consumption for each commodity (see Appendix S1). Lampung, Special Region of/*Daerah Istimewa* (DI) Yogyakarta, and Bali, were chosen for tobacco, SSBs, and alcoholic beverages, respectively.

### Analytical approach and stakeholders’ identification

We used a problem-driven Political Economic Analysis (PEA) adapted from Overseas Development Institute (ODI) framework(14, 15, 43) to guide our analysis. This framework offers flexibility in analyzing specific policies across a wide spectrum of issues(44, 45). Figure 1 below illustrates our adoption of the problem-driven PEA framework modified from Carriedo et al., 2021 (15). We adopted the framework by repeatedly aligning it with our problem statement and findings. We identified the current policy circumstances as a challenge for the health tax implementation at sub-national level in Indonesia. The structural issues include the social context and institutional arrangements, while agency issues encompass stakeholders and incentives. In our analysis of both structural and agency issues, we concentrated on organizations and institutions, including their roles, relationships, and interactions in the implementation of health taxation policies at the sub-national government level. Ultimately, through this analysis, we identified opportunities and barriers and proposed policy actions to be taken.

**Figure 1.**
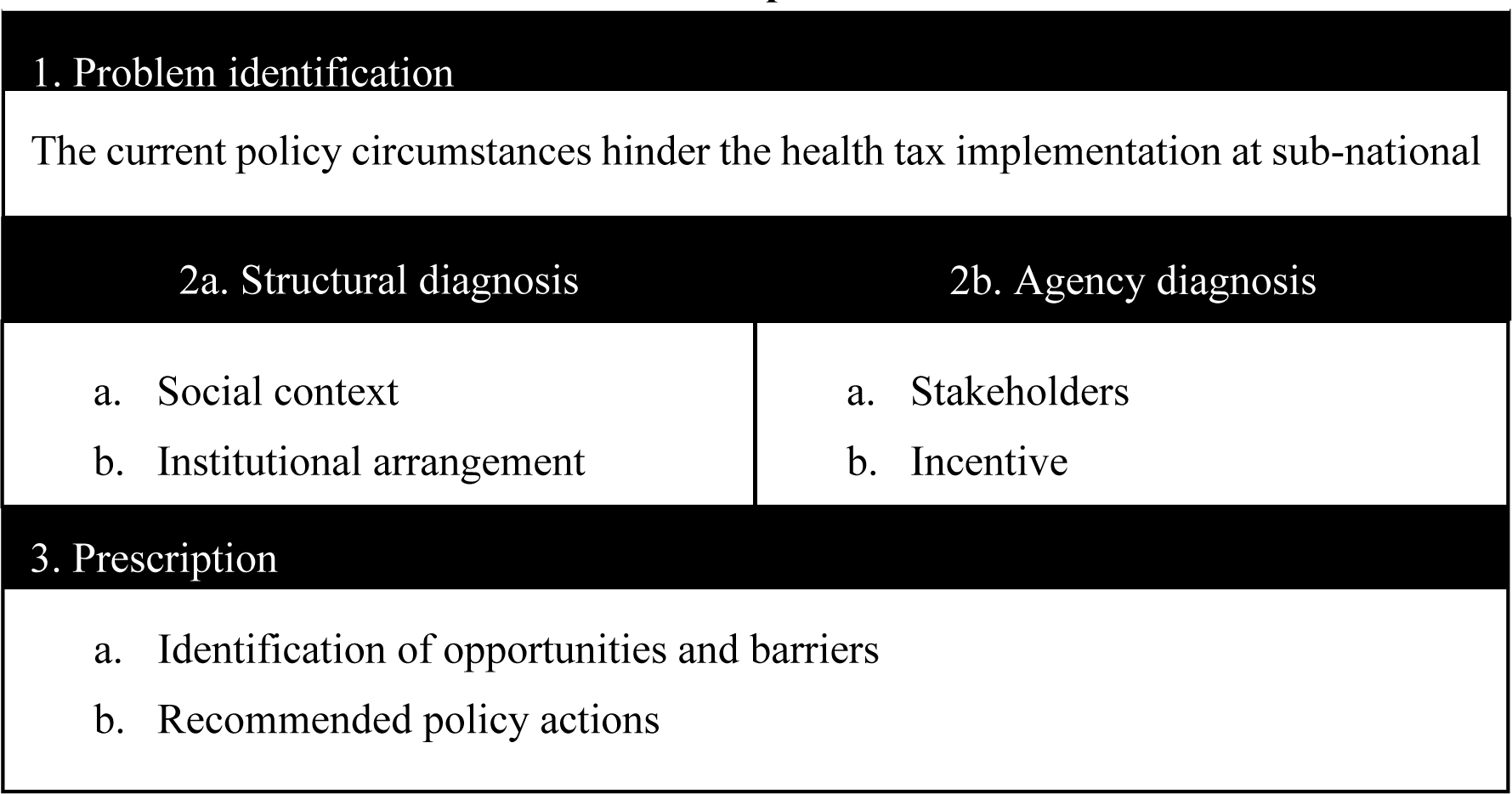
Problem-Driven Political Economy Analysis Approach, Adopted from Overseas Development Institute

We identified three relevant categories of participants. The first category is governmental institutions, which have in-depth information about governmental progress and cross-sectoral coordination and relationships. For each province, we recruited local government’s officer who are in charge of: (i) public finance; (ii) public health; (iii) human capital development; and (iv) development planning.

The second group comprises non-governmental organizations (NGOs) or experts. The information gained from these groups can serve as a counterbalance to the governmental institutions. For each province, the NGOs/experts recruited were from: (i) public health or economic experts from public/private higher education institution; (ii) local public health or economic NGOs; and (iii) local faith- or religious-based organizations.

Finally, the third group consists of consumers to better understand the social context and confirm the local government’s effort in increasing consumer awareness. Exploring consumers’ opinions also helped us gain more information from their perspective. The eligibility criteria for consumers to participate were those above the age of consent (18 years old) who regularly consume the researched commodity. Institutional stakeholders were recruited through close invitations, while individual (consumer) participants were recruited through open online invitations.

### Data collection

Semi-structured (see Appendix S2 for guiding questions) focus group discussions (FGD) were our primary source of data. This was selected based on our desire to explore a range of views, while observing the social interaction between participants(46). We conducted four separate FGDs in each province for each category of participants (see Table 1). Specifically for consumers, we organized FGDs for those aged 30 and above separately from those aged 18 to 30 to understand different behaviour among different age groups. In total, we conducted twelve FGDs between July and September 2022. Each FGD lasted approximately 120 minutes, accommodated a maximum of 15 participants, and arranged in a U-Shaped room setting. To ensure that participants understood the study’s context, each FGD began with a presentation by the researcher. All FGD activities were audio recorded and complemented with researchers’ notes. We reached data saturation at the last FGD and no additional data were collected.

**Table 1.** The Division of Role in Health Tax Management between National and Sub-National Governments in Indonesia.

| Product Category | Regulation | Description | Responsibility |
| --- | --- | --- | --- |
| Tobacco | Law No. 39/2007 on Excise Tax (as has been replaced by Law No. 7/2021 on Tax Harmonization) | Article 4: Any tobacco product (produced locally or imported) is subject to an excise tax. | • National government: determine the tax rate and tax collection. |
|  |  | Article 66A: Any excise tax collected from locally | • National government: disburse DBHCHT to |
|  |  | produced tobacco product is to be shared with the producing provinces through tobacco excise sharing fund (DBHCHT). | the provinces based on the share of their production.<br>• Sub-national government: manage and allocate the DBHCHT following the mandate of Ministry of Finance (MoF). |
|  | Law No. 28/2009 on Regional Tax and Retribution (as has been repealed by Law No. 1/2022) | Article 33: 10% of the excise tax on tobacco is to be dedicated for local tobacco tax. | • National government: collection and disbursement to the local government based on population size (MoF).<br>• Sub-national government: manage and allocate the local tobacco tax following the law mandate. |
| Alcoholic Beverages | Law No. 39/2007 on Excise Tax (as has been replaced by Law No. 7/2021 on Tax Harmonization) | Article 4: Ethyl alcohol and any beverages containing ethyl alcohol (produced locally or imported) is subject to an excise tax. | • National government: determine the tax rate and tax collection. |
| Sugar-Sweetened Beverages (SSBs) | N/A | A regulatory bill has been proposed since 2019; delayed implementation. | N/A; current regulatory draft is under MoF. |

**Table 1.**
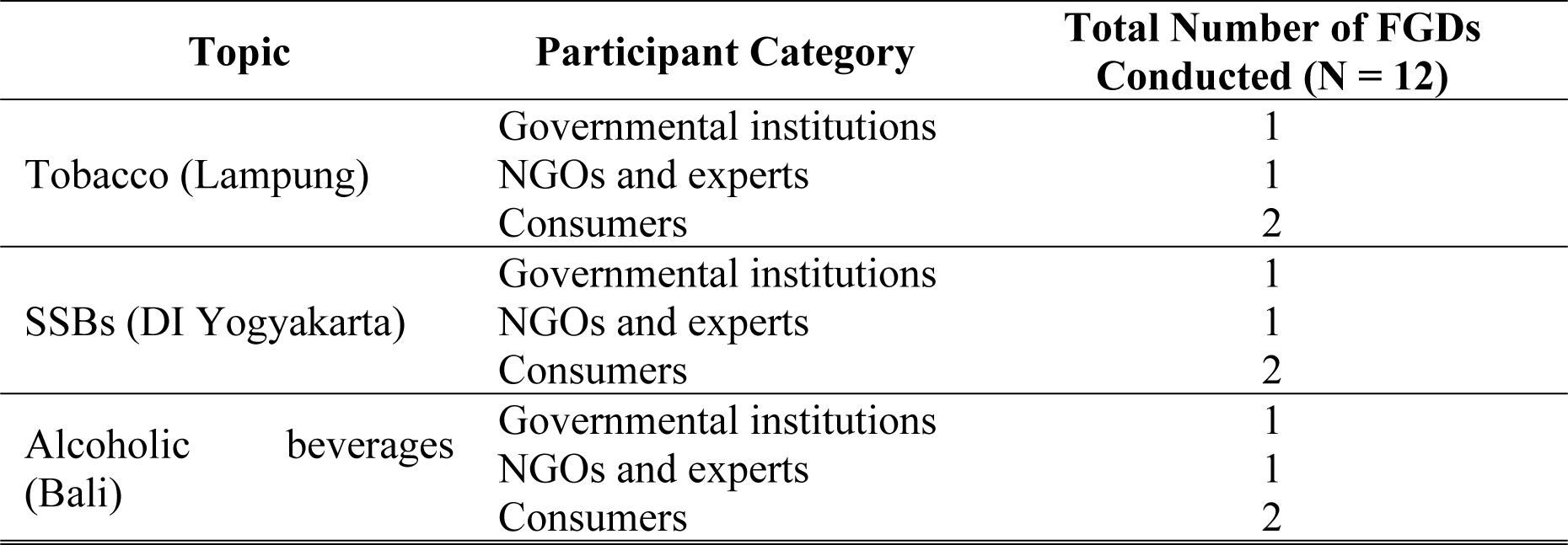
Summary of Focus Group Discussions (FGDs) Conducted.

| Topic | Participant Category | Total Number of FGDs Conducted (N = 12) |
| --- | --- | --- |
| Tobacco (Lampung) | Governmental institutions | 1 |
|  | NGOs and experts | 1 |
|  | Consumers | 2 |
| SSBs (DI Yogyakarta) | Governmental institutions | 1 |
|  | NGOs and experts | 1 |
|  | Consumers | 2 |
| Alcoholic beverages (Bali) | Governmental institutions | 1 |
|  | NGOs and experts | 1 |
|  | Consumers | 2 |

### Ethical consideration

This study received approval from the Ethical Review Board of Universitas Katolik Indonesia Atmajaya (Approval No. 00061/III/PPPE.PM.10.05/07/2022) and adhered to the Declaration of Helsinki(47). Prior to their participation, all individuals signed an informed consent, with the understanding that any subsequent publications arising from the study would not contain personal identifiers. To ensure the preservation of confidentiality, throughout this manuscript, participants are denoted by assigned pseudonyms.

### Data analysis

We employed framework analysis(48, 49) using the transcribed verbatim from all the audio records of the FGD. We constructed a matrix to display the participants’ arguments against the PEA framework, from which we derived keywords to align with the themes and sub-themes outlined in the PEA for each commodity category (see Appendix S3), adopted from Rodríguez et al., 2023 (14). A cross-member checking was performed to ensure rigorous data analysis process. The matrix for each commodity was independently developed by at least two researchers (e.g., KPR, YBAP and AE for alcoholic beverages; NHW, AE, and N Adhani for tobacco; and N Amalia, NHW, EVA, and MGU for SSBs) based on the verbatim transcript. The matrix appropriateness for each theme, sub-theme, and categories was iteratively reviewed by all team members. Regular research meetings were attended by all researchers to critically assess our findings before reporting the results. Disagreements among members were discussed, resolved, and managed accordingly.

## Results

We utilized the PEA framework to structure our findings into three key sections. Firstly, we provide an analysis of the context surrounding the health tax policy in each province. This analysis encompasses both structural and agency-related factors. Secondly, we explore and elucidate the barriers and challenges that are anticipated in the implementation of the policy. Lastly, we present the recommendations proposed by stakeholders to tackle these identified issues. In the results section, we compare the characteristics attributed to each commodity, highlighting how these differences shape the circumstances of each province’s case.

### Structural diagnosis

Structural diagnosis concerns the structural factors that shapes the stakeholders’ behaviour(45). We identified two sub-themes under the domain of the structural diagnosis in our PEA, i.e., social context and institutional arrangement.

### Social context

The social context defines the factors that shape health systems. In our case selection, where we have chosen cases based on the highest consumption levels for each commodity, this is closely linked to the underlying factors contributing to high consumption in each province, to gain a deeper understanding of the necessary policy interventions. Our qualitative analysis, conducted across three provinces, has identified three recurring keywords or categories. Firstly, cultural factors emerged as a significant determinant. Our focus group discussions with consumers and NGOs/experts revealed that high consumption rates are predominantly influenced by cultural factors, in addition to price-related factors, which underscore the need for reinforcing health taxes. Remarkably, this pattern holds true across all provinces, despite variations in the specific commodity category. For example, in Lampung, tobacco consumption is deeply ingrained in local traditions:

> *“Tobacco is a cultural item in Lampung during wedding ceremonies.” (NGOs 1, Lampung)*

This also applies to alcoholic beverages. Alcohol consumption in Bali is common during celebrations, ceremonies, or religious activities:

> *“Most Balinese people have arak (distilled spirit, traditional alcoholic drinks commonly found in Bali) at home for ceremonies, and there are numerous traditional alcoholic beverage producers in Bali. Therefore, owning alcohol in Bali is common. However, it is not consumed regularly; it is more of a by-*occasion*” (NGOs 1, Bali)*

Finally, in DI Yogyakarta, like other provinces, the consumption of sweetened foods and beverages appears to be commonplace due to the inclusion of sugary ingredients in many traditional food and beverage items. All three products are easily accessible at nearby stores and come at a affordable price, starting as low as IDR 500 (=USD 0.03):

> *“A sachet beverage can be as cheap as Rp* 500 *per pack, relatively inexpensive compared to daily expenses” (Consumer 1, DI Yogyakarta)*

A second aspect of the social context is consumer awareness. Our findings indicate that there may be varying levels of consumer awareness regarding regulated and unregulated commodities. In the case of regulated items such as tobacco and alcoholic beverages, despite high consumption rates, consumers are generally aware of the harm associated with these products, although many remain unaware of the specific purpose of taxing them. Conversely, in the case of SSBs, which are still unregulated, some consumers lack awareness of the long-term disadvantages of SSB consumption. As a common case in Asia, this is especially true for certain types of SSBs marketed as ‘healthy,’ such as dairy products or probiotic drinks, which mislead consumers into believing in their health benefits while remaining unaware of the sugar content:

> *“In my house, we usually stock up ready-to-*drink *tea and coffee and consume them up to three times a week. For probiotic drinks, it can be as much as three times a day” (Consumer 2, DI Yogyakarta)*

Consumers argued that due to current packaging, it is difficult to easily understand the nutritional content. Moreover, as there is no age limit for SSBs consumption, unlike tobacco and alcoholic beverages, parents make consumption decisions for children in their households, contributing to a relatively high SSBs consumption rate among individuals aged 3 and above. This common misconception among parents, exacerbates SSBs consumption as it can begin at an early age and lead to developing unhealthy dietary habits. Additionally, in the case of SSBs, advertising and packaging are recognized as two significant factors driving their purchase:

> *“The packaging is attractive, leading to* curiosity*. As in case for probiotic drinks, [I usually consume] for health” (Consumer 3, DI Yogyakarta)*

The third aspect is interlinked with the second factor in that consumer awareness shapes the social perception of a commodity. Regulated commodities such as tobacco and alcoholic beverages, which the government has labelled as harmful products, are also viewed negatively by consumers. Consumers have a stronger inclination to quit or at least reduce their consumption of these products. In contrast, SSBs, which have never been marketed negatively, are still seen as relatively normal and harmless commodities. Consequently, the implementation of health taxes is likely to be more challenging for SSBs compared to tobacco and alcoholic beverages.

### Institutional arrangement

In the institutional arrangement, we focus our discussion on the governmental and non-governmental institutions as well as the rules that shape power dynamics and outcomes, and the shared understanding between these institutions in adopting the national government policy. We identified two categories under the institutional arrangements dimension, e.g., ideology on the health tax objective and regulation and programs.

Firstly, the ideological perspective on the health tax objective concerns the shared understanding of policy objectives among each institution, with a primary focus on institutional participants. Majority of the institutions agreed that fiscal measure through health tax is needed for consumption control, particularly among specific demographic groups, including minors and lower-income individuals. Nevertheless, some disagreements emerged during these discussions. Institutions with a vested interest in industry and trade protection expressed reservations about the concept of health taxes. Their arguments centred on the potential disadvantages that health taxes could pose to local businesses operating at a micro-to-small scale. This concern is particularly relevant in the case of alcoholic beverages and SSBs, given that two provinces also host local businesses engaged in the production of these commodities:

> *“The sugary food and beverage industry is among the most* popular *in DI Yogyakarta. When Small and Microenterprises (SMEs) are asked to change their products, they will clearly refuse. Therefore, consumers are encouraged to control their intake.” (Dinkes, DI Yogyakarta)*

The second aspect of the institutional arrangement pertains to regulations and programs. The majority of sub-national level institutions argued that the authority for implementing fiscal measures to control NCDs lies solely with the national government. Sub-national governments primarily concentrate on the adoption of non-fiscal measures. This is particularly evident in the implementation of tobacco taxes, where the national government clearly defines the non-fiscal measures to be adopted by sub-national governments in accordance with Law No. 17/2023 on Health(50). In tobacco and alcoholic beverages cases, these measures include smoke-free area (specific for tobacco) and limit or ban advertising.

In the case of alcoholic beverages, although advertisements are more prominently visible in tourism areas, the sale of *arak* by micro enterprises remains prevalent. To address the potential impact of *arak* production, the Governor of Bali issued Governor Regulation No. 1/2020 on the Management of Balinese Fermented and/or Distilled Beverages(51). However, the orientation of this regulation is towards industry development, rather than protecting minors from alcohol consumption:

> *“There is Governor Regulation No. 1 of 2020, which aims to be a solution* for *marginalized Balinese alcoholic beverages [arak]. Many people in Bali used to consume imported beverages. Meanwhile, in Bali, we have our own distinctive Balinese alcoholic beverages, but due to regulations, they seem to be illegal,*

*causing traditional drinks to be undermined. [As a result] they are consumed less by our local community, which is unfair. Balinese beverages, distinct to Bali, have become illegal, while imported ones are considered legal.” (DinkopUKM, Bali)*

In the case of tobacco, even though Lampung has already established local regulations for Smoke-Free Areas (*Kawasan Tanpa Rokok*/KTR), tobacco advertisements can still be prominently observed along the main roads. This is compounded by the fact that microenterprises continue to sell tobacco to minors. Furthermore, the implementation of KTR itself remains notably weak:

> *“Many people continue to smoke in places where KTR have been* designated*. A significant portion of the population remains unaware of the KTR policy, and there are still people who smoke in hospital/health center areas.” (NGOs 1, Lampung)*

Regarding SSBs, while the fiscal measure has not yet been implemented, the non-fiscal measures that the sub-national government can take have not been clearly outlined. The initiative to raise issues related to limiting SSB consumption primarily came from public health government officials or NGOs/experts responsible for public health. Even in the governmental institutions responsible for the education sector or women and child empowerment, these initiatives have not been specifically adopted:

> *“Educational outreach to schools, especially in high schools and* vocational *schools, regarding health (the dangers of NCDs) has not specifically addressed SSBs.” (Disdik, DI Yogyakarta)*

### Agency Diagnosis

Agency diagnosis outlines power, incentives, and behavior(43). These three aspects are closely related to the stakeholders or actors who shape policy implementation. In our case, we are interested in discussing the main stakeholder’s power surrounding the implementation of health taxes at the sub-national level, as well as the factors influencing their behavior (i.e., motives) and their interactions. While sub-national governments do not have a role in tax collection itself, they play a strategic role in tax allocation and in assisting national government agencies in their areas (e.g., the Customs and Excise Office or *Kantor Perwakilan Bea dan Cukai*/KPBC). We have identified two main aspects of agency diagnosis in our case, namely stakeholders andincentives. See Figure 2 for a summary of the stakeholders, their decision-making power, and their motives that we have determined from our analysis.

**Figure 2.**
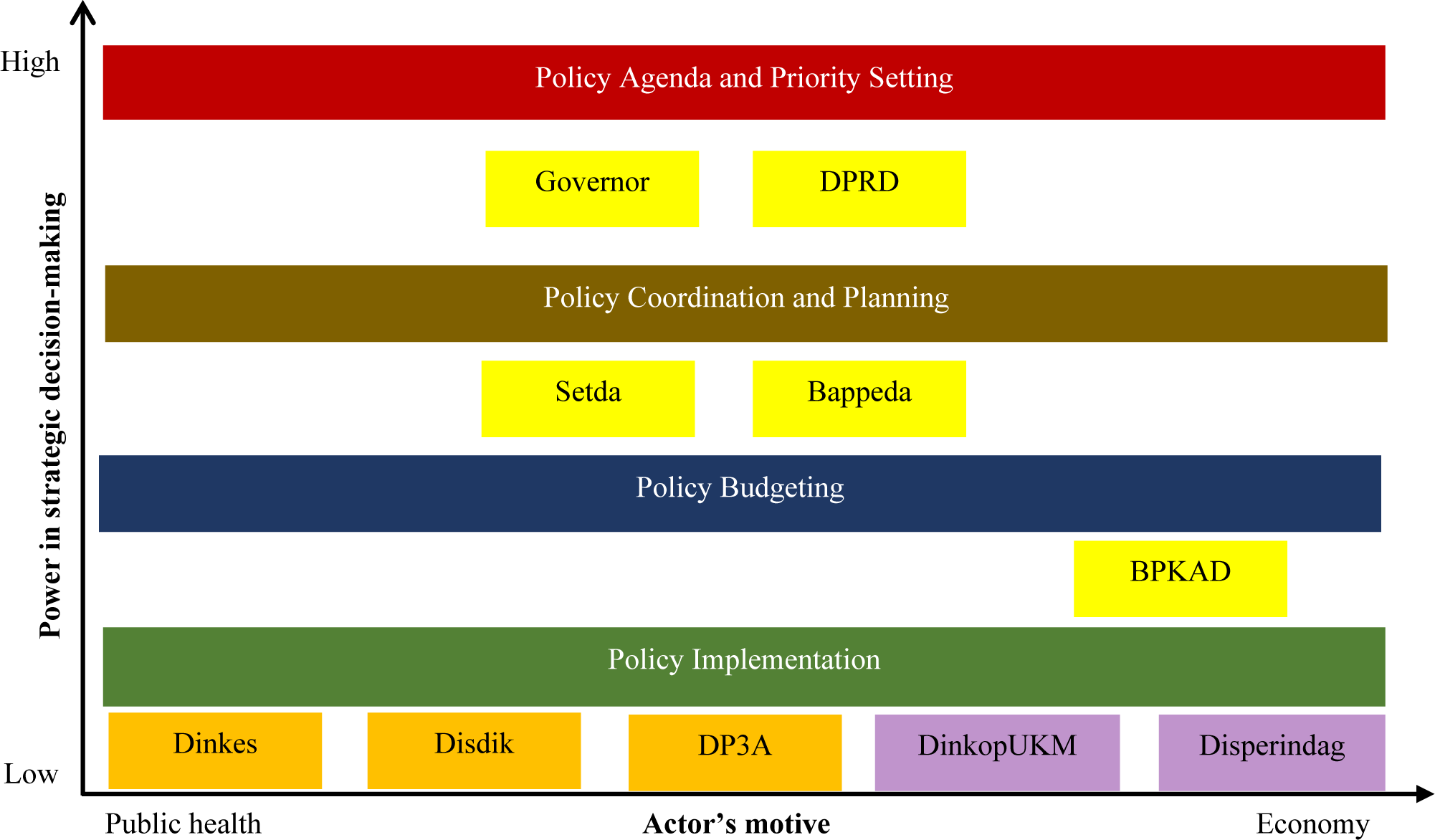
Key Actors Relationship in Supporting Health Tax Implementation at Sub-National Level in Indonesia

### Stakeholders

This sub-theme identifies key stakeholders in the implementation of sub-national health taxes, their roles, and their interactions (see Figure 2). Two topics have been derived from this sub-theme, i.e., key actors at the sub-national level and the power and relationships between actors at the sub-national level.

The first aspect comprises actors responsible for decision-making at the sub-national level. Based on our FGD findings, we have identified three categories (see Figure 2, the colour of each institution differs based on their roles) of actors at the sub-national level who can influence the adoption of health tax policies. While they may not be exactly the same across all provinces, they typically share similar sets of institutions/agencies. The first category (yellow coloured) includes actors responsible for strategic policy development and priority setting. At the provincial level, this includes the governor, the provincial regional legislative council (*Dewan Perwakilan Rakyat Daerah*/DPRD), the regional secretariat (*Sekretariat Daerah*/Setda), the regional agency of development planning (*Badan Perencanaan Pembangunan Daerah*/Bappeda), and the regional agency of finance and budgeting (*Badan Perencanaan Keuangan dan Anggaran Daerah*/BPKAD).

The second category comprises actors concerned with human capital development (orange coloured), including the regional department of health (*Dinas Kesehatan*/Dinkes), the department of education (*Dinas Pendidikan*/Disdik), and the department of child protection and women empowerment (*Dinas Pemberdayaan Perempuan dan Perlindungan Anak*/DP3A). Finally, the third category comprises actors in the economic sector (purple coloured), including the regional department of trade and industry (*Dinas Perdagangan dan Perindustrian*/Disperindag) and the department for cooperative and micro, small, and medium enterprises (*Dinas Koperasi dan UMKM*/DinkopUKM). In addition to these sub-national policy makers, there are actors without decision-making roles who interact with them: the local agency of the Customs and Excise Office—the representative of the Ministry of Finance in each region—assumes the responsibility for revenue collection and supervision at the local level, and the local agency of the Food and Drug Authority (*Badan Pengawas Obat dan Makanan*/BPOM), which exercises authority over nutritional value and safety in consumed products.

The second aspect is the power and relationships between the identified stakeholders. In this aspect, we identified four categories (see Figure 2). The four categories ranked from highest position in the strategic decision-making include the policy agenda and priority setting, policy coordination and planning, policy budgeting, and policy implementation, respectively.

The first category of actors holds higher positions in terms of regional decision-making compared to the other two categories. This is because they have the authority to set policy targets and priorities, which are then translated into policy planning and budget allocation by Bappeda in collaboration with BPKAD. Additionally, Setda and Bappeda coordinate and supervise intersectoral policy implementation. Actors in the human capital development sector and the economic sector mostly implement policies but can propose programs for prioritization before the budgeting process. They must provide evidence for why certain programs should be prioritized during the annual budgeting and planning process. Finally, the local KPBC does not hold a vertical position with other policymakers. However, in terms of implementing its supervisory role (i.e., eradicating illicit trade), it can collaborate with the sub-national government.

### Motives

This sub-theme pertains to the factors that shape the behavior of stakeholders. We have identified three main keywords that emerged from our analysis in this domain: the actors’ orientation/position towards public health and their incentive to support the health tax policy.

First is the actors’ orientation/position towards public health. To illustrate each institution’s objectives, we have created a spectrum of the actors’ orientation, ranging from public health motives to economic motives (see Figure 2). At the implementor level, Dinkes is the main actor with a public health orientation, along with other actors in human capital development.

However, they maintain a relatively neutral stance towards the health tax policy issue since it is not their core institutional motive, but they actively engage in public health advocacy:

> *“There are provincial and district programs for child-friendly areas with* indicators *that include a smoking ban. There are child forums throughout Indonesia that have actively campaigned against smoking with active outreach in schools. DP3A has also conducted anti-smoking campaigns at the neighborhood level, and there are regional regulations supporting smoke-free child-friendly areas. Disdik needs to be involved in raising awareness about the dangers of smoking.” (DP3A, Lampung)*.

While at the right hindside, for Disperindag and DinkopUKM, the orientation is more towards economy, in this case industry protection:

> *“As supporters of MSMEs, we agree because this excise tax applies to large* manufacturers*. Therefore, it can enable UMKMs to compete with large manufacturers.” (DinkopUKM, DI Yogyakarta)*

As for the coordinating bodies, BPKAD’s position leans more towards the economic aspect since their motive is to collect revenue for the regional government. We position Bappeda and Setda in a more neutral role since they both house units for human capital policy and economic policy development and primarily function as coordinators:

> *“This tax serves as an instrument to reduce consumption or circulation. I agree that this tax* should *increase, especially if there is revenue-sharing. The revenue-sharing from the tax can be used for SMEs in the alcoholic beverage industry and for awareness campaigns to improve the understanding that alcoholic beverages are detrimental to health.” (Bappeda, Bali)*

Lastly, political actors like the governor and DPRD are also in a neutral position due to their political motives, which can change with each election year.

Considering each actor’s motive, we can also identify their incentives for supporting and prioritizing the health tax policy. For Dinkes, their main performance indicator focuses on enhancing public health status, making them staunch supporters of adopting the health tax. In addition to the potential reduction in unhealthy consumption, this move offers substantial public health benefits. Furthermore, the public health sector anticipates receiving allocations from the tobacco tax and expects similar outcomes for other types of health taxes:

> *“I agree with the health tax and revenue-sharing that can be used to improve the quality and the* healthcare *sector.” (Dinkes, Bali)*

As for DP3A and Disdik, their support is somewhat conditional, hinging on the expectation that health tax revenue will be allocated to programs that bolster the implementation of health taxes. These programs could include initiatives to raise public awareness about unhealthy consumption in educational institutions or to aid women entrepreneurs in developing healthier product alternatives. These conditions are especially relevant in the case of SSBs, where there are currently no regulations in place to curb consumption, and public awareness is lacking:

> *“For health and for the education of children, women, families in rural areas, and SMEs. Because many of the* supported *families also produce juices and beverages, so they can have better awareness [on the harm of SSBs].” (DP3A, DI Yogyakarta)*

Conversely, Disperindag and DinkopUKM are primarily driven by the motive of safeguarding the local industry. Consequently, their support for health taxes hinges on whether these measures will protect local businesses. For instance, if the tax structure is tailored to the size of the industry, if it contributes to industrial development with an export focus, or if, in the case of SSBs, it serves to educate local industries about adopting healthier alternatives, these factors can influence their support:

> *“It [health tax] can be used for education and development of MSMEs [in the sector].” (DinkopUKM, DI* Yogyakarta)

Regarding BPKAD, their support for the health tax is more pronounced when the tax revenue can be earmarked for local government revenue, mirroring the structure of the tobacco tax. In the case of Bappeda, they maintain a more neutral stance toward the incentive, as policy priorities are subject to a bottom-up discussion, beginning with sector-specific priorities and later being communicated and decided upon by higher-ranking government entities such as Setda, the governor, and the DPRD.

### Way Forward

In this subsection, we delve into the main barriers and opportunities associated with the implementation of health tax policies at the sub-national level. Recommendations for addressing these barriers and optimizing opportunities will be discussed in the subsequent section.

#### Barriers

After conducting the structural and agency diagnosis, we can now identify potential barriers that may impede the implementation of health tax policies at the sub-national level. Three main barriers have been identified: a lack of awareness, distrust towards governmental institutions, and the existence of product substitution.

First and foremost is the issue of awareness. As discussed previously in the social context, there remains a significant lack of awareness among consumers regarding the health risks associated with the consumption of certain products. This lack of awareness is particularly pronounced in the cases of tobacco and SSBs. Concerning tobacco, the early age at which consumers often initiate smoking, combined with the addictive nature of tobacco, presents a formidable challenge in curbing consumption through the imposition of health taxes. While alcoholic beverages are also considered addictive substances, we do not observe the same patterns of consumption:

> *“It is somewhat impossible that Bali has the highest alcohol consumption rate. Most of it is related to traditional and religious purposes (it should be emphasized whether the factors driving this high alcohol consumption are for traditional and religious ceremonies or for recreational purposes and regular consumption).” (NGOs 2, Bali)*

Regarding SSBs consumption, the public still lacks a negative perception of these products, unlike the clear aversion to tobacco and alcoholic beverages. Furthermore, consumers currently struggle with understanding how to interpret the nutritional information on packaging and determining a safe sugar intake limit. Consequently, shifting their perspective on these products and raising awareness about limiting their consumption poses a significant challenge.

Secondly, there is an issue of trust in governmental institutions. Both consumers and institutions share a lack of trust in the government’s intent behind imposing health taxes. Presently, health taxes are predominantly viewed as revenue-generation tools rather than instruments for controlling consumption:

> *“I want this tax to be clearly used for a specific purpose. Let’s not* allow *it to be used solely for new political targets just to increase revenue.” (Consumer 3, DI Yogyakarta)*

Hence, consumers and institutions driven by the motive of safeguarding local industries still harbor reservations about the necessity of implementing health taxes, fearing potential harm to the local economy. The absence of transparent policy communication at the sub-national level, particularly to consumers, regarding the public health benefits of health tax implementation exacerbates this barrier.

Finally, there’s the challenge of product substitution and non-commercial consumption. Consumer substitution toward illicit alternatives represents a potential loss for both public health and revenue generation:

> *“I disagree because if the tax is* high*, it can trigger illicit tobacco due to low supervision in Indonesia. So, it shouldn’t be increased continuously; its upper limit should be assessed. The funds for law enforcement need to be increased to improve supervision.” (Bappeda, Lampung)*

However, an additional challenge arises from the fact that health taxes are levied on the end product rather than on the main ingredients of those products. In contrast to alcohol, tobacco is only taxed at the final product stage. Consequently, we can still observe instances where consumers purchase tobacco and hand-roll their own cigarettes, a practice known as “linting dewe.” In this scenario, the government has little control over the trade of these self-rolled cigarettes, and there are no limits on the quantity of tobacco that can be used, in contrast to commercially marketed products where such limits exist, potentially posing a health risk to the public:

> *“I don’t agree [with the increase in tobacco tax]. If it’s raised, I will still buy, maybe reduce a bit, I can do ‘linting dewe’ or choose cheaper cigarettes, but still stick to stick my smoking habits.” (Consumer 3, Lampung)*

This is also a concern raised by consumers and NGOs/experts regarding the implementation of taxes on sugar-sweetened beverages (SSBs). Many small-scale SSB manufacturers are unaware of the sugar content in their products. This lack of awareness extends to homemade SSBs as well, where consumers, lacking an understanding of the potential health risks associated with excessive sugar consumption, like homemade soy milk, may add as much sugar as they desire without recognizing the potential health drawbacks:

> *“Homemade SSBs are actually more risky because* there *are no [limit] for the sugar doses.” (Consumer 6, DI Yogyakarta)*

As a result of health tax implementation, which can lead to higher product prices, consumers are increasingly inclined to consider more affordable alternatives, including homemade options.

#### Opportunities

Regarding opportunities, we have identified two primary avenues to bolster the implementation of health taxes at the sub-national level. The first opportunity lies in garnering public support. While the lack of consumer awareness presents a challenge, there are individuals with a heightened awareness of the adverse health effects associated with tobacco, alcohol, and sugar-sweetened beverages (SSBs) who wholeheartedly endorse the health tax policy, even if they are consumers themselves. Consumers tend to rally behind policies that promise health benefits rather than revenue generation motives:

> *“I agree, because it relates to the non-communicable disease (NCD) prevention program. Because not all of the population is educated yet. If, for example, the price increases, it will compete with healthier food and drinks that are relatively expensive. But it’s also necessary to raise the base ingredients of sweetened beverages. It should be supported by education. Previous policies should also be maintained to support the tax.” (Consumer 7, DI Yogyakarta)*

The second opportunity centres around engagement with non-governmental organizations (NGOs). At times, sub-national governments lack the initiative to enhance public awareness, potentially undermining the effectiveness of health tax implementation. Local NGOs and academic institutions exhibit greater proactiveness in advocacy efforts. This presents an opportunity because focusing solely on engaging NGOs and academics at the national level may limit outreach to broader segments of society, especially beyond the Java region.

## Discussion

Indonesia, as one of the most decentralized nations, has encountered challenges in exercising local government autonomy, particularly in implementing fiscal autonomy (revenue generation) and delays in adopting national government regulations(20, 22, 52). Recognizing the complex nature of health tax implementation not only horizontally among national actors but also vertically among local actors, we built upon our previous work, specifically focusing on national-level policy debates on health taxes(53). This extension to the sub-national level becomes imperative as limited fiscal resources lead sub-national governments to prioritize curative over preventive or promotive measures in public health(54). Therefore, promoting health taxes as an additional source of funding for preventive and promotive actions earmarked for relevant programs in each institution is likely to gain more support from sub-national governments, as our findings suggest. This insight is especially important as political work to-date on sub-national health taxes, is limited to a small set of research on SSBs taxes in the US(55). Our findings also align with previous studies emphasizing the importance of earmarking health taxes for public health rather than framing them as revenue-generation tools(56–59). Although, detailing the well-intention of the health tax generation invites counteractive arguments from anti-tax opposition (55).

The same principle applies to policy communication. In Indonesia, policies driven by economic motives seem less likely to garner public support, whereas those driven by public health motives have a better chance of gaining popularity. Although more mixed supports suggested that emphasising the revenue streams for wider purposes, such as education initiatives, appear to be more appealing to voters(55). While recent cross-country work in LMICs has painted a rather mixed view about which arguments resonate in which settings, our findings are consistent with previous studies, suggesting that communicating health taxes as an effective measure to combat NCDs is likely to be publicly acceptable(60–63). However, without providing a convincing moral case for health taxes that resonates with diverse sets of interests, these measures might face challenges in the face of competing arguments from opponents(64, 65). Therefore, generating evidence to counter arguments from industry proponents should be a part of the policy agenda, as public acceptability is likely to be higher when strong evidence, provided by a credible source, supports a compelling claim(65–67).

In addition to allocating funds for promotive and preventive activities, health tax implementation can be complemented by other fiscal measures that are more applicable for sub-national governments. This could include subsidies for healthier alternatives, incentives for cessation(68–70), and regulatory and fiscal support for industries producing healthy products (71). Furthermore, health taxes often lead industries to reformulate their products to avoid the tax (72, 73), which could be an important tax design consideration(74). Therefore, health tax revenue can also be allocated to enable sub-national governments to promote and ensure proper product reformulation for healthier alternatives by engaging local enterprises. This is consistent with our finding that institutions with industry protection motives support health taxes because they can benefit local enterprises, particularly for promoting healthier alternatives.

In addition to fiscal measures, non-fiscal measures are equally important. While the majority of past studies have shown a negative relationship between health taxes and unhealthy consumption or NCD prevalence(70, 75–80), anticipating consumers’ substitution with cheaper but equally harmful alternatives is important(68, 81). Therefore, increasing consumer awareness to consciously limit their consumption of unhealthy products should be a priority. This can be achieved through extensive efforts to promote a healthy lifestyle through mass campaigns, civil society engagement, and women’s empowerment(60, 61, 82–84). Our findings support the importance of optimizing the engagement of local NGOs and academics, as well as raising awareness among institutions interested in women’s empowerment and children(63), such as Disdik and DP3A.

Another critical non-fiscal measure that should be strongly enforced is a complete ban or strict limitations on advertising. While regulatory frameworks for tobacco advertising are in place, lack of local government commitment and weak surveillance systems hinder adoption at the sub-national level(22, 85). While we found no issues with alcohol advertising, our findings suggest potential disadvantages of advertising and packaging information for SSBs. Since there are currently no specific interventions for SSBs, past studies have suggested Front-of-Pack (FOP) or ‘traffic-light’ nutritional labeling (86–88). This should be accompanied by extensive consumer education to increase their understanding of nutritional labeling, as previous studies have noted a lack of comprehension among consumers(89). Furthermore, in the realm of law enforcement, the health tax can also be leveraged to impose more stringent penalties, enhance monitoring efforts, and provide counselling for sellers found illegally selling tobacco and alcohol to minors.(90–92).

Finally, given the cross-sectoral nature of health tax implementation, sub-national and national governments must share a common understanding of the policy objectives to avoid policy polarization. With health financing problems being one of the decentralization issues facing the healthcare sector in Indonesia(35), framing health taxes as a means of health financing will be crucial to avoid conflicting interests in tax allocation. This should be a clear priority during the planning process to ensure budget alignment and avoid implementation delays(14, 16, 93).

We propose three crucial recommendations for advancing the implementation of health taxes, applicable to both sub-national and national governments.

First and foremost is the aspect of policy communication, which holds paramount importance in gaining the trust and support of the public. When communicating the health tax policy, advocates should consider a wider range of arguments in support of health taxes, such as ones based on health impacts, as opposed to prioritizing just the revenue-generation potential of these taxes.

Secondly, the health tax earmarking should be a key consideration. Even though sub-national governments may not directly engage in health tax collection, they can actively participate in the allocation of health tax revenues. Although hard earmarking is subject to a debate, a soft earmarking which allows a more flexible funding priorities and aligns with decentralization, can garner greater public and sub-national government support for health taxes.

In addition to the health sector, as is currently the case with tobacco, some broader recommendations for funding allocation includes: (i) enhancing public awareness through preventive and promotive actions; (ii) supporting industrial development with an export orientation, especially for alcoholic beverages, which may have unique local specialties suitable for export; (iii) raising awareness among local small and micro-enterprises to encourage shifts toward healthier product alternatives, such as product reformulation (particularly for SSBs); (iv) providing subsidies for healthier alternatives, particularly targeting lower-income households; and (v) strengthening law enforcement, which includes eradicating the illicit trade of cigarettes and imposing fines on those who sell alcoholic beverages and tobacco to minors.

Lastly, non-fiscal measures are equally critical alongside fiscal ones. Some significant non-fiscal measures that deserve prioritization include: (i) implementing bans or limitations on advertising, an area currently under sub-national government authority. This restriction should also extend to SSBs. Furthermore, the national government should consider reinforcing the regulatory framework to encourage sub-national governments to adopt this policy; (ii) enforcing standardized packaging, similar to the tobacco industry, which features easily recognizable packaging with pictorial health warnings. We propose that this approach be applied to SSBs as well. The government should contemplate implementing mandatory Front-of-Pack or ‘traffic light’ labelling to mitigate consumer confusion; and (iii) launching extensive promotional campaigns, particularly for SSBs. In the case of tobacco, which has long been marketed as a ‘harmful’ product, similar efforts should be directed toward raising awareness about the health risks associated with SSB consumption.

To the best of our knowledge, this is the first attempt to analyze the political economy of health taxes for three different commodities (tobacco, alcoholic beverages, and SSBs) at the sub-national level in Indonesia. However, this study is subject to certain limitations. Firstly, it is constrained by its focus on specific provinces, potentially posing a threat to external validity. However, we took steps to mitigate this limitation by broadening our sample to encompass three provinces, each with different commodities under scrutiny. Additionally, the inclusion of multiple stakeholders from various sectors, including government, non-governmental organizations, and consumers, provides a more comprehensive perspective that may be applicable in other settings.

Secondly, our study did not involve elected policy actors such as governors or members of the DPRD. These individuals may offer valuable insights into the political economy of health taxes. Future research should consider expanding the pool of informants to include these elected officials, thereby enriching our understanding of the subject.

Lastly, our study’s scope is limited in terms of SSBs knowledge. This limitation arises because, at the time of our data collection, the tax on SSBs had not been implemented. Subsequent studies should consider conducting research specifically on SSBs after the implementation of relevant regulations to provide a more comprehensive analysis of the impact of health taxes on this particular commodity.

## Conclusion

This study aims to understand the political economy of health tax policy implementation at the sub-national level by critically exploring the case of Indonesia as one of the most decentralized countries. The findings of this study identify lessons learned for countries facing similar challenges of a complex decentralized decision-making process.

Sub-national governments, constrained by limited fiscal resources, tend to prioritize curative actions over preventive measures. To address this, promoting health taxes as a funding source for preventive initiatives is crucial, aligning with findings from limited research on sub-national health taxes. However, counteractive arguments from anti-tax opposition should be anticipated and addressed through comprehensive communication strategies.

Policy communication should emphasize a wide range of arguments, including health impacts, to gain public support. Earmarking health tax revenues, with a preference for soft earmarking, allows flexibility and aligns with decentralization principles, garnering greater support. Recommendations include prioritizing public awareness, supporting industrial development, and providing subsidies for healthier alternatives. Non-fiscal measures, such as advertising restrictions and standardized packaging, are crucial. The study, focusing on tobacco, alcoholic beverages, and SSBs at the sub-national level, acknowledges limitations and suggests future research directions, emphasizing the need for a comprehensive understanding of health taxes’ political economy.

## Data Availability

All original data is accessible to the public by contacting the corresponding author. The original data for editorial and reviewer teams has been enclosed within the system (as a supporting information)

## Acknowledgement

The authors would like to thank Robert Marten and Kaung Suu Lwin at the Alliance for Health Policy and Systems Research (a WHO-hosted partnership), as well as Adam Koon for the comments and insights during the preparation of this manuscript. This research study is part of a series of analytical country case studies to better understand the political economy of advancing health taxes supported by the Alliance for Health Policy and Systems Research, in collaboration with WHO Departments and the Inter-Agency Working Group on Health Taxes.

---

**Appendix S1.** The Prevalence of Tobacco, Alcoholic Beverages, and Sugar-Sweetened Beverages (SSBs) in Indonesia by Province, 2018*.

| Province | Tobacco<sup>a</sup> | Alcoholic Beverages<sup>b</sup> | Sugar-Sweetened Beverages (SSBs)<sup>c</sup> |
| --- | --- | --- | --- |
| Aceh | 24.0 | 0.4 | 59.81 |
| North Sumatera | 22.4 | 5.5 | 58.51 |
| West Sumatera | 26.9 | 1.5 | 60.63 |
| Riau | 25.3 | 2.2 | 62.39 |
| Jambi | 21.5 | 0.7 | 54.46 |
| South Sumatera | 25.3 | 1.2 | 63.82 |
| Bengkulu | 27.8 | 1.7 | 55.81 |
| Lampung | 28.1 | 1.8 | 65.95 |
| Bangka Belitung | 24.8 | 3.0 | 57.26 |
| Kepulauan Riau | 22.3 | 3.3 | 59.58 |
| DKI Jakarta | 22.9 | 3.0 | 61.72 |
| West Java | 27.1 | 1.7 | 63.91 |
| Central Java | 23.2 | 1.9 | 68.82 |
| DI Yogyakarta | 19.5 | 3.2 | 71.01 |
| East Java | 23.9 | 1.8 | 57.00 |
| Banten | 26.8 | 2.1 | 61.46 |
| Bali | 18.9 | 14.0 | 42.90 |
| West Nusa Tenggara | 26.3 | 3.4 | 51.73 |
| East Nusa Tenggara | 19.0 | 15.6 | 53.60 |
| West Kalimantan | 23.5 | 7.5 | 59.05 |
| Central Kalimantan | 24.3 | 4.2 | 64.57 |
| South Kalimantan | 20.6 | 1.3 | 70.82 |
| East Kalimantan | 21.9 | 2.9 | 61.57 |
| North Kalimantan | 22.5 | 4.5 | 67.53 |
| North Sulawesi | 23.5 | 16.0 | 60.94 |
| Central Sulawesi | 26.1 | 8.8 | 58.67 |
| South Sulawesi | 22.0 | 6.3 | 56.29 |
| Southeast Sulawesi | 22.3 | 6.2 | 52.66 |
| Gorontalo | 27.4 | 11.3 | 56.16 |
| West Sulawesi | 21.8 | 4.1 | 58.25 |
| Maluku | 22.1 | 11.1 | 62.06 |
| North Maluku | 23.9 | 7.2 | 59.40 |
| West Papua | 22.1 | 7.5 | 59.18 |
| Papua | 18.8 | 4.9 | 53.45 |

\*Source: Basic Health Research Report, Ministry of Health of Republic of Indonesia, 2018
<sup>a</sup>Population aged 10+, consume daily
<sup>b</sup>Population aged 10+, based on past month consumption
<sup>c</sup>Population aged 3+, consume at least once per day.

### Appendix S2 Focus Group Guide and Informed Consent

#### Informed Consent Form

“Health Taxes Implementation Challenges in Indonesia: Case Study of Three Provinces” I hereby declare that:

a. My participation is voluntary, and I may withdraw from this Focus Group Discussion (FGD) at any time without the obligation to provide reasons. Should I choose to withdraw, all related data will be promptly erased and will not be utilized further;
b. I agree that the information I provide within this FGD will exclusively serve research purposes and the subsequent publication of research findings;
c. I consent to the understanding that my personal data will not be employed or disclosed in any form within this research;
d. I agree to the recording of the FGD process and the documentation of salient points presented therein;
e. I have perused and comprehended the information pertaining to this research. I have had the opportunity to pose questions and have received satisfactory and adequate responses concerning this research.

In light of the aforementioned considerations, I willingly consent to participate as an informant in this research, fully cognizant and free form any coercion or duress.

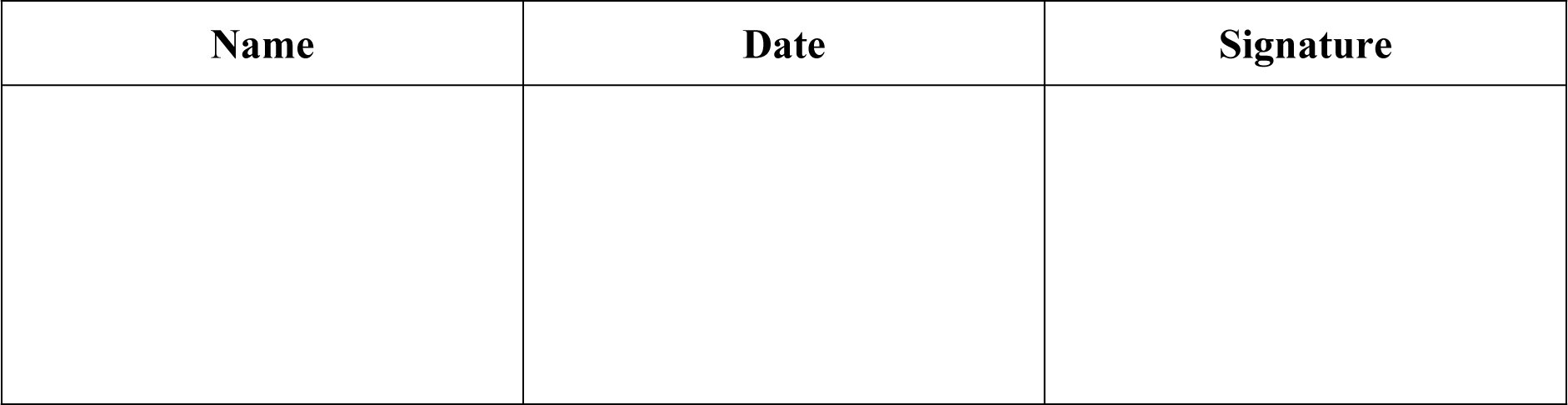

#### Focus Group Discussion

“Health Taxes Implementation Challenges in Indonesia: Case Study of Three Provinces”

##### Introduction

Good morning/afternoon, Ladies and Gentlemen. We are a team of researchers from Universitas Indonesia. My name is, and I will serve as the moderator for this Focus Group Discussion (FGD) session. Firstly, we’d like to thank you for your participation in this FGD. The session today is expected to last no more than 120 minutes, and each participant will have the opportunity to share their opinions on each question.

We want to reassure you that this FGD session will be fully audio-recorded, and all information gathered during and after this FGD will be used exclusively for research purposes. The confidentiality of your personal information is of utmost importance to us. We have provided you with an informed consent form, ensuring that your participation remains completely anonymous. We will not disclose any personal identifiers or the identity of your institution.

Before we dive into the session, we will begin with a brief presentation outlining the objectives and context of this research.

##### List of questions (for institutions)

1. What are the primary public health issues on your institution’s agenda? Probe: are there any related to diseases caused by the consumption of tobacco/alcohol/SSBs?
2. How does your institution position itself in efforts to control non-communicable diseases caused by the consumption of tobacco/alcohol/SSBs? Probe: health motive, economic motive
3. Can you please describe your understanding of a tobacco/alcohol/SSBs taxes?
4. Can you briefly explain your institution’s opinion on the current implementation of tobacco/alcohol/SSBs taxes in your province?
5. How would you evaluate the current implementation of tobacco/alcohol/SSBs taxes implementation in your province?

##### List of questions (for consumers)

1. Can you describe your consumption patterns for cigarettes/alcohol/SSBs? Probe: total daily consumption, percentage of your budget spent, the type/brand you consumemost frequently.
2. Why do you think it’s important for you to regularly consume cigarettes/alcohol/SSBs? Probe: taste, any events, emotions, or situations associated with consumption.
3. In your opinion, how does the consumption of cigarettes/alcohol/SSBs in your family and community compare to five years ago? Check: has it increased or decreased
4. Can you describe your feelings when you watch/see advertisements for cigarettes/alcohol/SSBs? Probe: negative/positive emotions, purchase intention.
5. Can you describe your feelings when you watch/read news or public health information in the media about cigarettes/alcohol/SSBs? Probe: negative/positive emotions, awareness, sense of control.
6. What is your understanding of cigarette/alcohol/SSBs taxes?
7. If the price of cigarettes/alcohol/SSBs were to increase due to taxes, how would you respond? Investigate: agree or disagree, why
8. In your opinion, what should the government do to reduce alcohol consumption? Investigate: public awareness campaigns, clear regulations, etc.

**Appendix S3.** Abstraction Matrix for Framework Analysis using Political Economy Approach.

| Theme | STRUCTURAL DIAGNOSIS |  | AGENCY DIAGNOSIS |  | WAY FORWARD |  |  |
| --- | --- | --- | --- | --- | --- | --- | --- |
| Sub-theme | Social context | Institutional arrangement | Stakeholders | Incentive | Barriers | Opportunities | Recommendation |
| **Definition** | *(Contextual factors that impact the problem)* | *(Institutions and rules that shape power dynamics and outcomes as well as the shared understanding between institutions in adopting the national government policy)* | *(Identification of stakeholders, their role, and their interaction)* | *(Factors that shape stakeholders' behavior)* | *(Barriers that hinder the policy implementation)* | *(Opportunities that could support the policy implementation)* | *(Recommended actions to overcome barriers and optimizing opportunities)* |
| **Keywords** | Cultural factors | Ideology on the health tax objective | Key actors | Actors orientation/position towards public health | Lack of awareness | Public support | Policy communication |
|  | Consumers' awareness | Current regulations and programs | Power and relationships between actors | Incentive to support and prioritize the policy | Trust towards governmental institution | NGO engagement | Earmarking |
|  | Social construct of the commodity attribute |  |  |  | Product substitution and not-for-commercial consumption |  | Non-fiscal measure |

